# Dementia and Alzheimer’s disease in former contact sports participants: population-based cohort study, systematic review, and meta-analysis

**DOI:** 10.1101/2022.05.24.22275500

**Authors:** G. David Batty, Philipp Frank, Urho M. Kujala, Seppo J. Sarna, Carlos A. Valencia-Hernández, Jaakko Kaprio

**Affiliations:** Department of Epidemiology and Public Health, University College London, London, UK; Faculty of Sport and Health Sciences, University of Jyväskylä, Jyväskylä, Finland; Department of Public Health, University of Helsinki, Helsinki, Finland; National Heart & Lung Institute, Imperial College London, London, UK; Institute for Molecular Medicine FIMM, University of Helsinki, Helsinki, Finland

## Abstract

**Importance:** There is uncertainty regarding the long-term risk of dementia in individuals with a history of participation in sports characterised by repetitive head impact, and whether the occurrence of this disease differs between former amateur and professional athletes.

**Objective:** To quantify the dementia risk in former athletes with a background in contact sports using new cohort study data incorporated into a meta-analysis.

**Data sources and study selection:** The cohort study comprised 2005 male retired athletes who had competed internationally for Finland (1920-1965) and a general population comparison group (N=1386). For the systematic review, we searched PubMed and Embase from their inception to August 23 2022, including cohort studies published in English that reported standard estimates of association and variance.

**Data extraction and synthesis:** Studies were screened and results extracted independently by two authors. Study-specific estimates were aggregated using random-effect meta-analysis. An adapted Cochrane Risk of Bias Tool was used to assess study quality.

**Main outcomes and measures:** The primary outcomes were dementia and Alzheimer’s disease.

**Results:** The systematic review identified 827 potentially eligible published articles, of which 7 met the inclusion criteria. Incorporating the new results from the Finnish cohort study with those from the systematic review revealed that former boxers had higher rates of dementia (2 studies: summary risk ratio 3.14 [95% CI 1.72, 5.74], I^2^=34%) and Alzheimer’s disease (2 studies: 3.07 [1.01, 9.38], I^2^=55%), as did retired soccer players (3 studies of dementia: 2.78 [1.69, 4.59], I^2^=86%; 2 studies of Alzheimer’s disease: 3.22 [1.34, 7.75], I^2^=81%). While the pooled estimate for dementia in retired American football players was less convincing (4 studies: 1.63 [0.76, 3.49], I^2^ =75%), disease occurrence in onetime professionals was markedly higher (2.96 [1.66, 5.30]) than amateurs in whom there was no association (0.90 [0.52, 1.56]). There was also a risk differential for retired professional (3.61 [2.92, 4.45]) and amateur (1.60 [1.11, 2.30]) soccer players.

**Conclusion and relevance:** Based on studies exclusively comprising men, former participants in contact sports subsequently experienced poorer brain health, and there was a suggestion that retired professionals had the greatest risk.

**Key points:** *Question:* Do former participants in contact sports have a greater risk of dementia than the general population?

*Findings:* Compared with general population controls, retired male participants from the contact sports of boxing, soccer, and American football appeared to have an elevated risk of dementia at follow-up. For soccer and American football, the risk in former professionals was higher than erstwhile amateurs.

*Meaning:* Retired male contact sports participants seem to subsequently experience poorer brain health.

## Introduction

Estimates of the present and future burden of dementia for individuals, societies, and health care systems are clear: there are currently around 60 million cases globally and, owing to demographic growth of the ageing population, predictions are that this is likely to triple by 2050.^1^ Unsuccessful trials of drug treatment to alter the course of the disease brings into sharp focus the need to identify modifiable risk factors for the primary prevention of dementia,^2^ and head injury has recently been advanced in this context.^3^

Head impact occurs on a spectrum of severity. In people who experience head trauma serious enough to warrant hospitalisation, typically resulting from motor vehicle accidents or falls, there is evidence from large scale cohort studies of the general population of an approximate doubling in the rate of later dementia relative to the unexposed,^4-7^ although these are not universal findings.^8^ There is also an indication of a dose-response association, whereby less severe head injury has intermediate level risk.^9^ These observations raise concerns that lower intensity repetitive head impact, characteristic of participation in contact sports, may have implications for the occurrence of dementia.^10^

Although autopsy investigations reveal evidence of chronic traumatic encephalopathy, a neurodegenerative disease, among select samples of former athletes from American football,^11^ soccer,^12^ and boxing,^13^ whether participation in these sports is associated with a later risk of dementia is uncertain. In former American football players, for instance, there is a suggestion of elevated rates of dementia relative to the general population,^14^ while other studies report no difference in the occurrence of this neurodegenerative disorder across these groups.^15^ Elsewhere, dementia risk was raised amongst retired professional soccer players,^16,17^ yet a small cohort of onetime amateur boxers had the same rates as unexposed controls.^18^

To address this paucity of evidence, we first utilise new data from a well-characterised cohort study of men drawn from an array of elite amateur athletic backgrounds which include the contact sports of soccer, wrestling, and boxing, who had long-term follow-up for incident dementia. We then aggregate these results within a meta-analysis based on a systematic review of results from all published cohort studies. Although there are several recent systematic reviews of dementia in former contact sports participants,^19-21^ we are not aware of any quantitative synthesis of these findings, including a comparison of disease occurrence in retired amateur and professional athletes who have different levels of historical exposure to head impact.

## Methods

### Cohort of Finnish former elite athletes

This cohort study was initiated in 1978 to examine the relationship between participation in different elite-level sports and longevity.^22,23^ The sample was generated from 2613 Finnish male former elite amateur athletes (640 Olympians) who competed internationally between 1920 and 1965 (supplemental figure 1). Identified using a combination of sports yearbooks, association records, and periodicals, these individuals represented a diverse array of sporting backgrounds. Data collection was approved by the ethics committee of the Hospital Districts of Helsinki and Uusimaa, and all participants consented. The reporting of this cohort study conforms to the Strengthening the Reporting of Observational Studies in Epidemiology (STROBE) Statement of guidelines for reporting observational studies.^24^

#### Derivation of exposed and unexposed groups

Based on the occurrence of head injury,^25-27^ the included sports were denoted as contact (soccer, boxing, wrestling, ice hockey, and basketball) or non-contact (track and field, cross-country skiing, and weight-lifting). Individual contact sports were then disaggregated as the numbers of dementia cases at follow-up allowed. Thus, separate analyses were possible for former soccer players, boxers, and wrestlers (non-professional or freestyle/Greco-Roman), while retired athletes from the sports of ice hockey and basketball players were combined into a ‘other’ contact sports category. A general population-based (unexposed) comparison group comprising age-equivalent healthy men was generated using records from a population-wide medical examination routinely administered to all Finnish males at around 20 years of age as part of national service (N=1712). Participants in non-contact sports represented a further group unexposed to head impact.

#### Assessment of covariates

Data on covariates were extracted from two sources. The presence of diabetes, hypertension, and coronary heart disease was derived from linkage of study members to a national drug treatment register. Additionally, in 1985, surviving study members and population controls (N=2851; 66% of the original cohort) were mailed a self-completion questionnaire (N=1917; response 67%) (supplemental figure 1) with enquiries regarding health behaviours (e.g., smoking, alcohol intake), physical stature, and weight. Questionnaire data in combination with those extracted from the Finnish Central Population Registry were used to generate a variable for longest held job, our indicator of socioeconomic status.^28^

#### Ascertainment of dementia

Health surveillance began upon initiation of nationwide health registries in 1970 when the average age of the athlete group was 45.4 years (controls 44.3 years). Study members were linked to: death records, including cause; hospitalisation records, including cause; and the Finnish Drug Prescription Register for medications specifically used to treat Alzheimer’s disease (see supplemental table 1 for International Classification of Disease and medication codes). Using these data, we derived three dementia outcomes for use in the present analyses: dementia, Alzheimer’s disease, and non-Alzheimer’s dementia.

### Systematic review and meta-analysis

#### Search strategy and study selection

This PROSPERO-registered (CRD42022352780) systematic review and meta-analysis was reported in accordance with Preferred Reporting Items for Systematic reviews and Meta-Analyses (PRISMA)^29^ and Meta-analysis Of Observational Studies in Epidemiology (MOOSE)^30^ guidelines. We identified relevant literature by searching PubMed (Medline) and Embase databases between their inception and August 23 2022. We used combinations of free text and controlled terms in 2 categories (supplemental box 1): the exposure (i.e., specific sports such as boxing, soccer, martial arts, and rugby), and the outcome (i.e., dementia and its subtypes such as Alzheimer’s disease and vascular dementia). We also scrutinised the reference sections of retrieved articles for additional publications.

We included a published paper if it fulfilled the following criteria: utilised a cohort study; the diagnosis of dementia outcomes was made by a healthcare professional as part of a medical examination in either a research or clinical setting (self-reports of physician diagnosis and cognitive tests did not qualify); reported standard estimates of association (e.g., relative risk, odds ratios, hazard ratios) and variance (e.g., confidence interval, standard error); appeared in a peer-reviewed journal; and was published in English. Two authors (GDB and PF) independently screened the identified records first by title, then abstract, and, if necessary, the full paper. Any discrepancies were resolved on discussion.

#### Extraction of results and assessment of study quality

Where available, a range of characteristics were extracted from each publication, including the name of the lead author, publication year, country of sample population, number of participants exposed/unexposed, number of events and type, and effects estimates for both minimally- and multivariable-adjusted results. Study authors were contacted when clarification was required.

We used an adapted version of the Cochrane Risk of Bias Tool to assess study quality.^31^ This comprised seven domains of appraisal, including the comprehensiveness of the exposure, confounding variables, outcome ascertainment, and adequacy of the follow-up (supplemental table 2). With a maximum of 4 points for each domain (total 28), we regarded the quality of the study as high if the total score was at least 21, moderate if the score ranged between 14 and 20, and low if it was <14.

#### Statistical analyses

In individual-participant analyses of the Finnish cohort study, after exclusion of study members owing to record-linkage failure and death prior to the beginning of follow-up, the analytical sample comprised 3,391 men (2005 former athletes, 1386 population controls). Event surveillance was from 1^st^ January 1970 until the occurrence of a dementia event or the end of surveillance (December 31, 2015) – whichever came first. Having ascertained that the proportional hazards assumption had not been violated, we used Cox regression to compute hazard ratios with accompanying 95% confidence intervals to summarise the relationship of a background in contact sports with the risk of dementia.^32^ Age was used as the time covariate.

For the meta-analysis, we pooled the results from the analyses of raw data in the Finnish cohort alongside published study-specific estimates using a random effects meta-analysis,^33^ an approach which incorporates the heterogeneity of effects in the computation of their aggregation. An I^2^ statistic was computed to summarise the heterogeneity in estimates across studies. All analyses were computed using Stata 15 (StataCorp, College Station, TX).

## Results

### Finnish cohort study

In analyses of the Finnish cohort, a mean of 30.6 years of disease surveillance (range 1-46 years) in the analytical sample of 3391 men, gave rise to 406 cases of dementia (265 cases of Alzheimer’s disease). There was little difference in the magnitude of the hazard ratios in the univariate and multivariate models (table 1), so we present the results for the latter here. Former boxers experienced a more than three-fold risk of dementia (hazard ratio 3.60 [95% CI 2.46, 5.28]) relative to the general population comparison group. While risk was also elevated in retired wrestlers (1.51 [0.98, 2.34]) and soccer players (1.55 [1.00, 2.41]), statistical significance was not apparent. The risk of dementia in erstwhile athletes from ‘other’ contact sports (ice hockey and basketball) and those from non-contact sports such as track and field was not significantly different to the general population controls. Alzheimer’s disease was most strongly related to a history of contact sports, whereby boxing was associated with a quadrupling in risk (4.10 [2.55, 6.61]), and the rate in wrestlers (2.11 [1.28, 3,48]) and soccer players (2.07 [1.23, 3.46]) was around double that of general population controls. The risk of non-Alzheimer’s dementia was also raised in retired boxers (2.62; 1.68, 4.11) but not convincingly so in former soccer players (1.23 [0.74, 2.06]) nor wrestlers (1.22 [0.73, 2.03]) (supplemental table 3).

**Table 1.** Hazard ratios (95% confidence intervals) for the association of former participation in contact sports with dementias and Alzheimer’s disease: Finnish cohort study.

|  | n/N | Dementia |  | Alzheimer's disease |  |
| --- | --- | --- | --- | --- | --- |
|  |  | Age-adjusted<br>(406/3391) | Age-, SES-, co-<br>morbidity, and<br>lifestyle factors-<br>adjusted<br>(288/1916) | Age-adjusted<br>(265 / 3391) | Age-, SES-, co-<br>morbidity, and<br>lifestyle factors-<br>adjusted<br>(188/1917) |
| Boxing | 64 / 230 | 3.68<br>(2.72, 4.99) | 3.60<br>(2.46, 5.28) | 4.45<br>(3.07, 6.45) | 4.10<br>(2.55, 6.61) |
| Wrestling | 38 / 247 | 1.31<br>(0.91, 1.89) | 1.51<br>(0.98, 2.34) | 1.90<br>(1.24, 2.90) | 2.11<br>(1.28, 3.48) |
| Soccer | 44 / 248 | 1.80<br>(1.27, 2.54) | 1.55<br>(1.00, 2.41) | 2.43<br>(1.62, 3.64) | 2.07<br>(1.23, 3.46) |
| Other contact sports | 18 / 236 | 0.74<br>(0.45, 1.22) | 0.73<br>(0.41, 1.30) | 0.85<br>(0.46, 1.56) | 0.80<br>(0.39, 1.62) |
| Non-contact sports | 122 / 1,044 | 0.88<br>(0.68, 1.13) | 0.80<br>(0.58, 1.11) | 0.89<br>(0.64, 1.23) | 0.84<br>(0.55, 1.28) |
| General population (controls) | 120 / 1,386 | 1.0 (ref) | 1.0 (ref) | 1.0 (ref) | 1.0 (ref) |

n=number of events; N=group size. Number of events and group size according to individual sports and are for age-adjusted analyses of dementia. SES, socioeconomic status. Co-morbidities are diabetes, hypertension, and coronary heart disease. Lifestyle factors are alcohol intake, passing out due to alcohol, body mass index, and smoking status.

We conducted some sensitivity analyses. First, given the maturity of this cohort, competing risk bias may arise because the occurrence of a non-dementia event precludes occurrence of our primary outcome of interest. We therefore subjected Alzheimer’s disease to a competing risks analysis based on the most common cause of ill-health herein, cardiovascular disease, and our findings were unchanged (supplemental table 4). Second, given the amateur era in which study members competed internationally, it is plausible that, if sufficiently skilled, they could have participated at elite level in more than one of the sports linked to dementia in our analyses. Mutual control for each of the three sports related to dementia in earlier analyses did not, however, change the overall pattern of association (supplemental table 5). Lastly, owing to missing data, the more complex statistical models comprise fewer study members. We therefore recomputed the main effects in a non-missing dataset and results were essentially the same (supplemental table 6).

### Systematic review and meta-analysis

The systematic review identified 827 potentially eligible published articles of which 7 met our inclusion criteria (figure 1). Of these, four cohort studies captured dementia cases in retired American football players^14,15,34,35^ (none featured Alzheimer’s disease); there were two such studies of former soccer players^16,17^ (one with Alzheimer’s disease,^16^ one with non-Alzheimer’s dementia^16^); and one of retired boxers^18^ (one with Alzheimer’s disease^18^) (table 2). One study was field-based^18^ and the remainder were generated from linkage of administrative records. All retrieved studies exclusively comprised men, and were a mix of former amateurs^15,18,34^ and professionals.^14,16,17,35^ Comparison groups were typically the general population, with two utilising a non-collision sports group.^15,35^ The studies of erstwhile American football players were unsurprisingly based on samples from the USA,^14,15,34,35^ whereas former boxers were drawn from Wales,^18^ and the soccer players from Scotland^16^ and France.^17^ When reported, the number of events in the retired athletes ranged from 5^18^ to 180^16^ for dementia; 2^18^ to 64^16^ for Alzheimer’s disease; and for the single study reporting non-Alzheimer’s dementia the count was 64.^16^ Of the seven retrieved studies, only one^16^ was evaluated as being of high quality, with the remainder classified as moderate (supplemental table 2).

**Figure 1.**
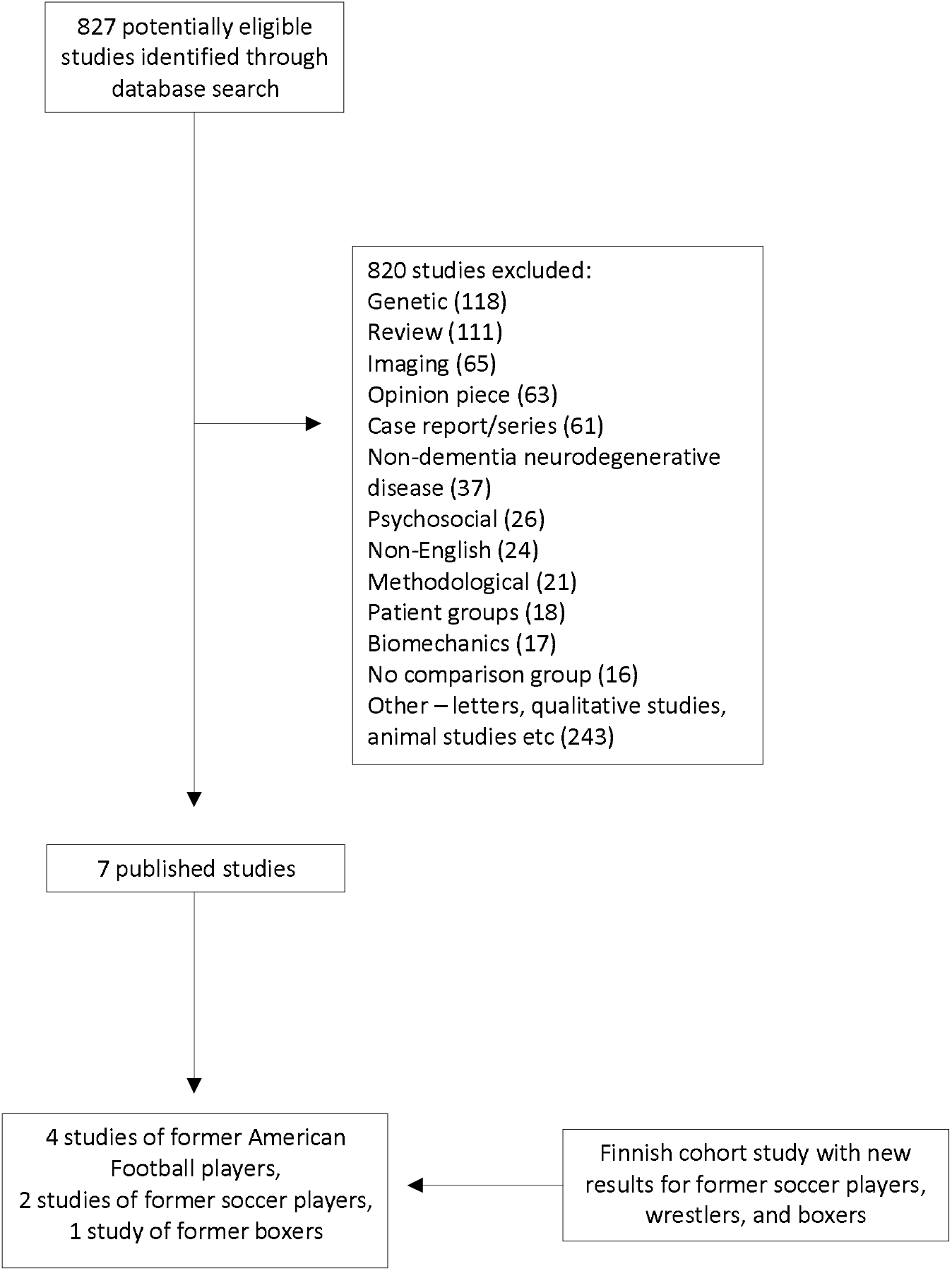
Study selection: Systematic review.

**Table 2.**
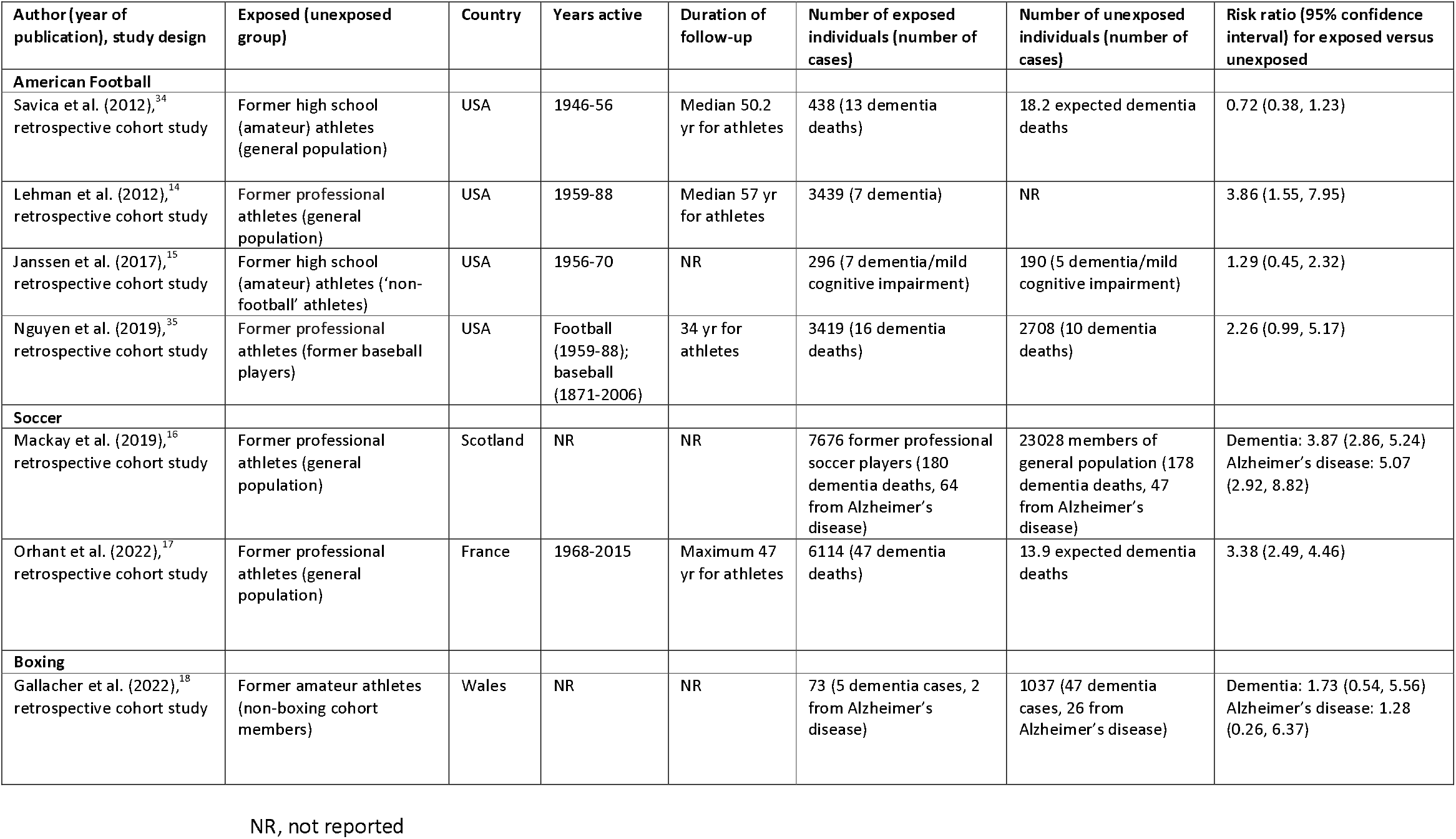
Former participation in contact sports and dementia: Characteristics of included studies.

In the meta-analysis, we pooled results from these published studies alongside the new findings from the Finnish cohort. On combining effects estimates for all contact sports, there was around a doubling of the risk of dementia (summary risk ratio 1.91 [95% CI 1.30, 2.82], I^2^=87%) and Alzheimer’s disease (2.30 [1.32 to 4.00], I^2^=83%) relative to general population comparison groups.

In analyses of specific sports, former soccer players (figure 2) had elevated rates of dementia (2.78 [1.69, 4.59]), Alzheimer’s disease (3.22 [1.34, 7.75]), and non-Alzheimer’s dementia (2.32 [1.04, 5.16]). Confidence intervals around these estimates were wide, however, indicating low precision, and there was a high degree of cross-study heterogeneity in both analyses (I^2^>81%). In the studies of former professional soccer athletes, there was a suggestion of a higher risk of dementia (3.61 [2.92, 4.45]) relative to the single cohort of retired amateur players (1.60 [1.11, 2.30]).

**Figure 2.**
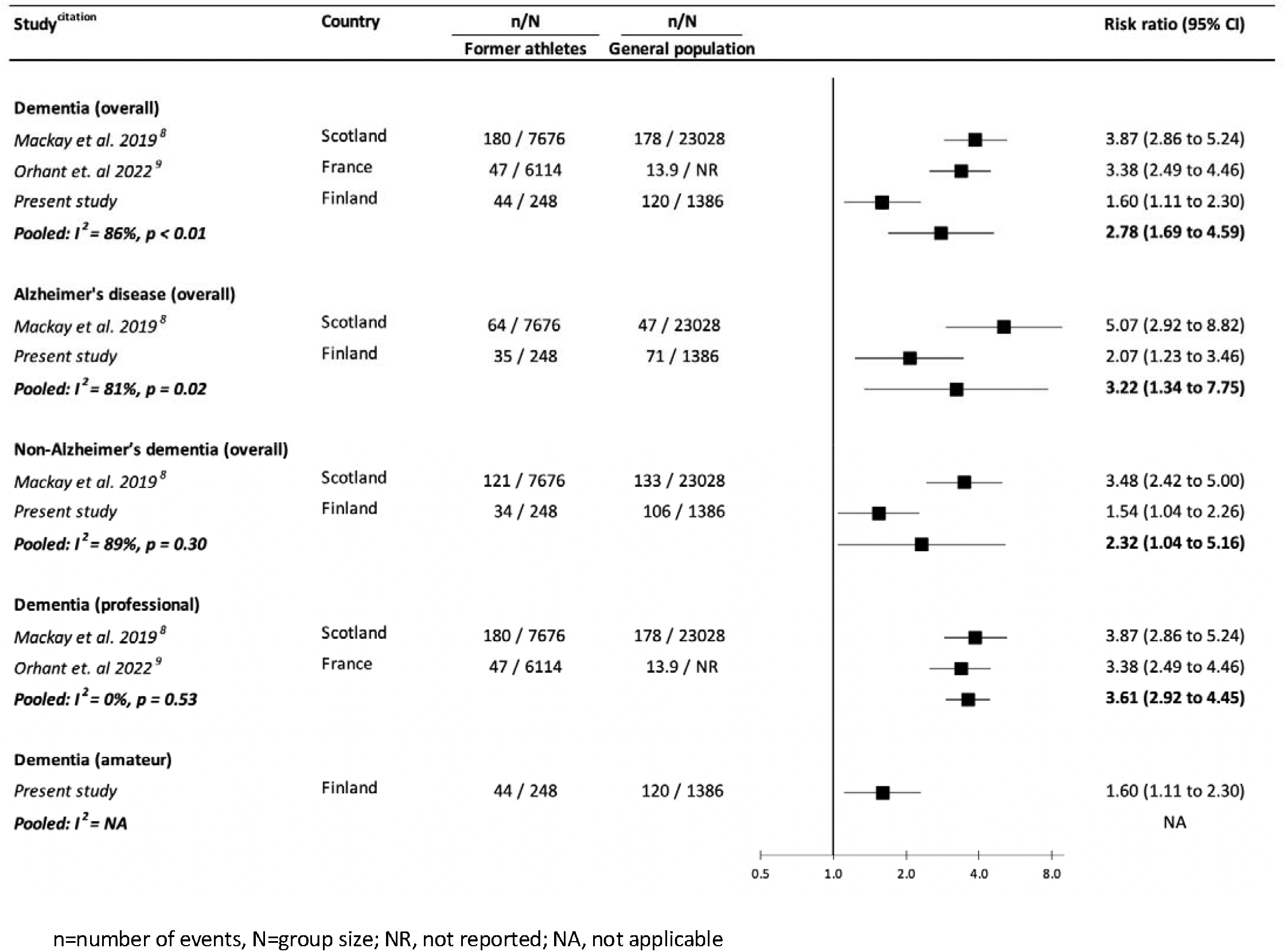
Risk ratios (95% confidence intervals) for the association of former participation in professional and amateur soccer with dementia and Alzheimer’s disease: Meta-analysis.

The overall results for the American football studies (figure 3) revealed an unconvincingly raised risk of dementia in former participants, the confidence interval for which included unity (1.63 [0.76, 3.49], I^2^=75%). Again, however, a differential in dementia risk according to past playing level was also evident, whereby an association was evident for retired professionals (2.96 [1.66, 5.30]) but not in onetime amateurs whose only exposure was in high school (0.90 [0.52, 1.56]). In one of the American football studies,^15^ the ‘unexposed’ group of non-football players in fact comprised some wrestlers who, in analyses of the Finnish cohort study herein, had a modestly elevated post-retirement risk of dementia. This could have had the impact of narrowing the risk differential; however, dropping the study from the meta-analysis did not alter our conclusions.

**Figure 3.**
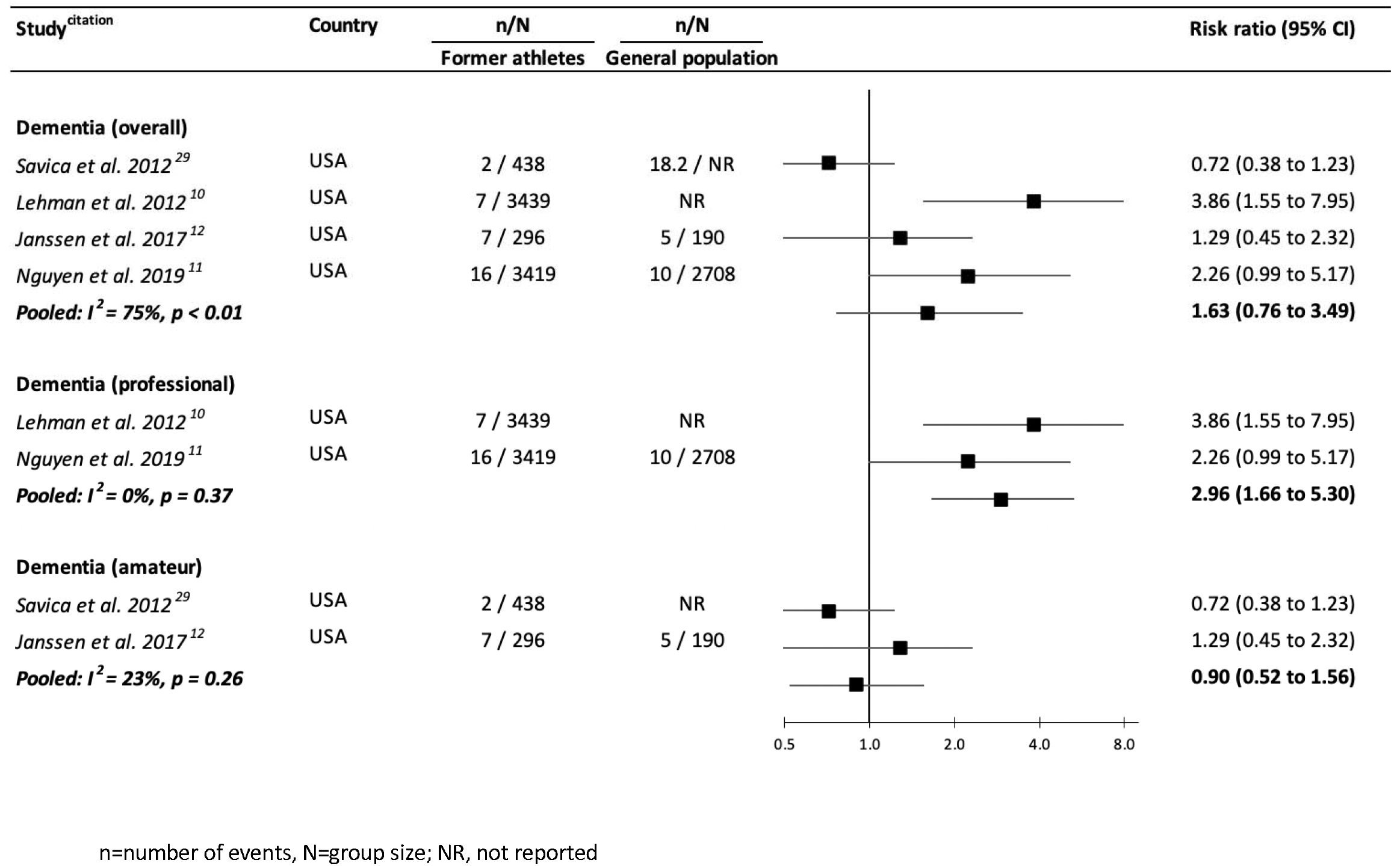
Risk ratios (95% confidence intervals) for the association of former participation in professional and amateur American football with dementia: Meta-analysis.

Lastly, in the analyses of boxers (figure 4), all of whom competed on an amateur basis, in aggregate, former athletes experienced a greater risk of both dementia (3.14 [1.72, 5.74]) and Alzheimer’s disease (3.07 [1.01, 9.38]). There was a moderate level of heterogeneity across these two studies (I^2^≤55%).

**Figure 4.**
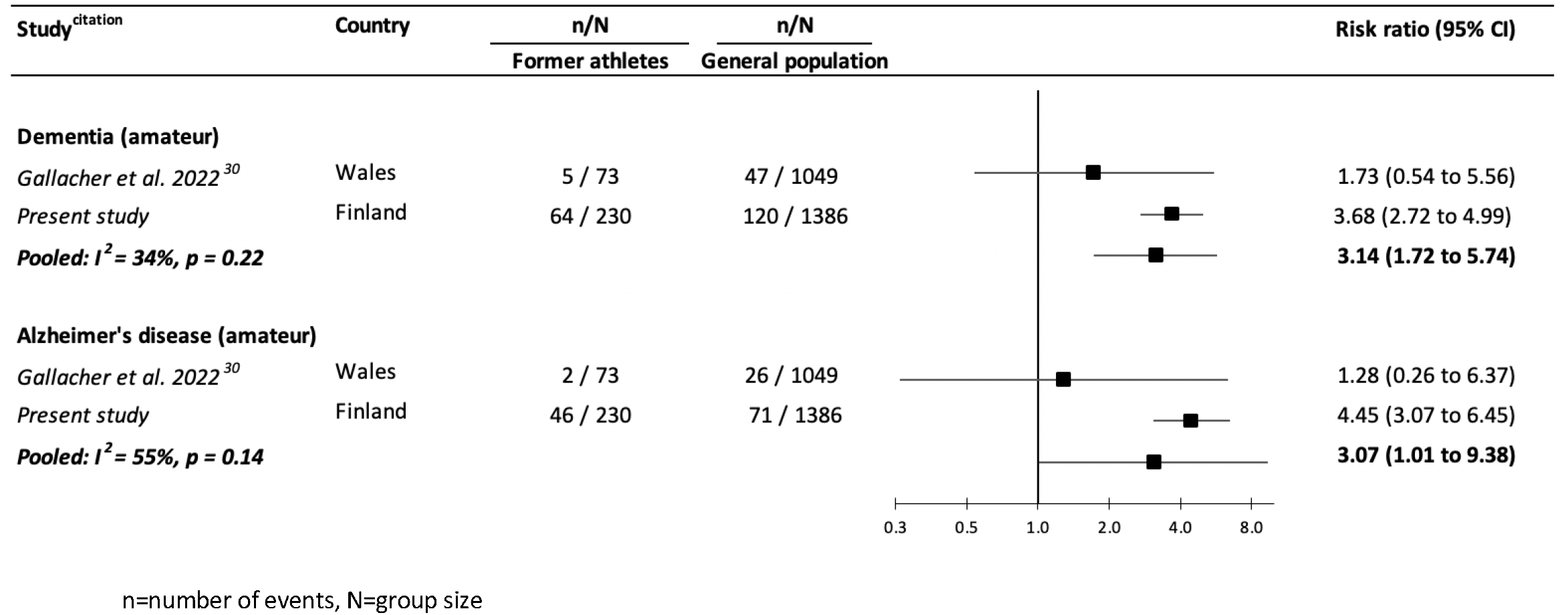
Risk ratios (95% confidence intervals) for the association of former participation in amateur boxing with dementia and Alzheimer’s disease: Meta-analysis.

## Discussion

In the present study, we quantitatively aggregated results from new analyses of a Finnish cohort study with those from the extant literature. With the caveats that the evidence base is modest in scale and confined to men, relative to general population comparison groups, we found elevated rates of dementia in former participants from the contact sports of soccer and boxing. While studies of retired American football players did not collectively reveal a convincing association with dementia, those from professional backgrounds experienced markedly higher risk than amateurs who were only exposed in high school. This differential in disease risk between former professionals and amateurs was also seen for former soccer athletes. In partial support of the finding of lower dementia rates in former high school football athletes, at autopsy, former professionals and college players had a markedly higher prevalence of chronic traumatic encephalopathy, a neurodegenerative disorder.^11^

Another salient finding of the present meta-analysis was the general pattern of dementia risk despite the contrasting head impact profiles of the included sports. That is, concussions are most common in wrestling and boxing,^26 27^ and result largely from person-on-person contact – in boxing, for instance, the primary objective is to render the opponent unconscious. In soccer, however, the insult is more likely to be sub-concussive and attributable to contact with the ball when attempting to redirect it using the head. In within cohort analyses of former soccer players featured in the present meta-analysis,^16^ those who occupied outfield positions in their careers and were therefore more likely to ‘head’ the ball, had higher rates of dementia at follow-up, including Alzheimer’s, relative to goalkeepers who rarely head the ball.^36^

### Plausible mechanisms

Most of the studies included in the present meta-analysis had a modest collection of confounding factors, however, in the analyses of the well-characterised Finnish cohort, controlling for an array of risk factors had little impact on the magnitude of the effects. This apparent independent influence of contact sports participation therefore raises the question of potential mechanisms. It is plausible that athleticism and neurodegenerative disorders share selected gene variants although empirical data are currently lacking. For those sports played in outdoor environments, it has been speculated that exposure to pesticides might raise dementia risk,^37,38^ though this would not apply to the indoor sports of boxing and wrestling where dementia risk was highest. Sport-specific explanations include, for soccer, the suggestion that heading produces an acute if temporary lowering of cognitive function^39^ while potentially more permanent structural changes to the brain are evident upon imaging.^40^ Specific mechanisms of risk in boxing and American football are less clear. That global participation in soccer – estimated at more than a quarter of a million by its governing body^41^ – is seemingly the highest of any sport, and programmes of wrestling and American football are long-established in many colleges,^42^ means that, if indeed causal, a link between a background in these activities and dementia may have public health significance.

### Study strengths and limitations

The present study has its strengths, including being the first quantitative synthesis of dementia risk in former participants from a range of contact sports, and one that incorporates new cohort study data. It is not, however, without its limitations. First, all retrieved studies, plus the Finnish cohort, exclusively sample men. While we can think of no reason for the present results not applying to women given that other risk factors predict dementia equally well in sex-specific analyses,^43,44^ clearly, empirical studies are required. Second, the findings of a meta-analysis are only as strong as the methodological quality of the studies it includes, and all data herein were observational with the large majority of moderate quality. With conventional trials in this field being unviable ethically and perhaps logistically, an advance on current evidence may be the use of natural experiments. These could include the impact on dementia risk pre- and post-introduction of compulsory protective equipment such as change in the composition of the soccer ball from leather to plastic which resulted in lower weight (1986-present), or the introduction of head gear in amateur boxing (1984-2016). This quasi-experimental approach has been used elsewhere in dementia research by examining the role of mandated increases in school leaving age as a proxy for improved education.^45^ Lastly, none of the studies have data on actual head impacts; instead, sporting background was used as a proxy. There is empirical evidence, however, of a higher occurrence of head trauma in contact sports groups versus control populations.^15^

In conclusion, based on a modest collection of studies of moderate quality, former participants in contact sports experienced poorer brain health at follow-up, and this was particularly evident for retired professionals with the highest accumulative exposure to head impacts. Whether these findings are generalisable to women is unclear given the dearth of data.

## Data Availability

Bona fide researchers should contact the authors to discuss the availability of raw data.

## Acknowledgement

The preparation of this manuscript received no direct funding. GDB is supported by the UK Medical Research Council (MR/P023444/1) and the US National Institute on Aging (1R56AG052519- 01; 1R01AG052519-01A1); PF by the UK Economic and Social Research Council & Biotechnology and Biological Sciences Research Council (Soc-B Centre for Doctoral Training); and JK by the Academy of Finland Centre of Excellence in Complex Disease Genetics (336823).

