## Supplemental material for "Dementia and Alzheimer’s disease in former contact sports participants: population-based cohort study, systematic review, and meta-analysis" (from another version of this preprint: version 3, 2022-09-22)

**Supplemental Figure 1. Flow of individuals into the analytical sample: Finnish cohort study**

4325 men (2613 elite athletes competitive 1920-1965, and 1712 population-based controls) identified

Excluded: 379 athletes & 276 controls dying before start of follow-up in 1970; and 279 untraceable

406 dementia cases

265 Alzheimer’s disease cases

333 non-Alzheimer’s disease dementia cases

289 dementia cases

188 Alzheimer’s disease cases

228 non-Alzheimer’s disease dementia cases

3391 surviving men (2005 former athletes, 1386 controls) with retrospective and prospective linkage to health records from 1970

2851 surviving study members mailed questionnaire in 1985 (N=1917, 67% response) with follow-up from Jan 1, 1986

**Supplemental Table 1. International Classification of Disease codes for study outcomes: Finnish cohort study**

| **Endpoint** | **ICD-8** | **ICD-9** | **ICD-10** |
| --- | --- | --- | --- |
| Dementia, not otherwise  specified | 290.0, 290.1, 290.19 | 290.0A, 294.1A, 331.0A,  331.1A, 331.2X, 331.8X, 437.8A | F01-F03;  G30; G31 |
| Alzheimer’s disease* | 290.1 | 331.0A | G30 |
| Non-Alzheimer’s dementias | 290.0, 290.19 | 290.0A, 294.1A, 331.1A, 331.2X, 331.8X, 437.8A | F01-F03; G31 |

*Anatomical Therapeutic Chemical codes for drugs to treat Alzheimer’s disease based on the Finnish Drug Prescription Register for

medications: N06DA02, donepezil; N06DA03, rivastigmine; N06DA04, galantamine; and N06DX01, memantine

**Supplemental Table 2. Assessment of study quality based on the Cochrane Risk of Bias Tool:**

**Systematic review**

|  | **I**  **Exposed and unexposed from same population** | **II Confidence in exposure assessment** | **III**  **Outcome absent at baseline** | **IV Comprehensive adjustments** | **V**  **Confidence in confounders assessment** | **VI Confidence in outcome assessment** | **VII Adequate follow-up** | **Total (/28)** |
| --- | --- | --- | --- | --- | --- | --- | --- | --- |
| **Soccer** |  |  |  |  |  |  |  |  |
| Mackay (2019) | 3 | 4 | 4 | 2 | 2 | 4 | 3 | 22 |
| Orhant (2022) | 3 | 3 | 4 | 1 | 1 | 1 | 1 | 14 |
| **Boxing** |  |  |  |  |  |  |  |  |
| Gallacher (2022) | 4 | 3 | 1 | 4 | 4 | 3 | 1 | 20 |
| **American Football** |  |  |  |  |  |  |  |  |
| Savica (2012) | 4 | 3 | 4 | 1 | 1 | 3 | 2 | 18 |
| Lehman (2012) | 2 | 4 | 3 | 1 | 1 | 3 | 1 | 15 |
| Janssen (2017) | 4 | 0 | 4 | 1 | 1 | 3 | 1 | 14 |
| Nguyen (2019) | 2 | 3 | 3 | 2 | 2 | 3 | 0 | 15 |

To assess study quality, we responded to the following enquiries: I. Was selection of exposed and non‐exposed cohorts drawn

from the same population? II. Can we be confident in the assessment of exposure? III. Can we be confident that the outcome of

interest was not present at the star of the study? IV. Did the statistical analysis adjust for the confounding variables? V. Can we be

confident in the assessment of the presence or absence of confounding factors? VI. Can we be confident in the assessment of outcome?

VII. Was the follow up of cohorts adequate? The studies were evaluated in relation to each question using 4 categories: “definitely yes”

(score 4), “probably/mostly yes” (3), “probably/mostly no” (2), and “definitely no” (1). Maximum of 4 points for each domain (total 28).

Quality of the study was denoted as being high if the total score was at least 21, moderate if the score ranged between 14 and 20,

and low if it was <14.

**Supplemental Table 3.** **Hazard ratios (95% confidence intervals) for the association of former participation**

**in contact sports with non-Alzheimer’s dementia: Finnish cohort study**

|  | **Number of events / Number at risk** | **Age-adjustment**  (333/3,391) | **Age-, SES-, co-morbidity-, and lifestyle factors-adjusted**  (227/1916) |
| --- | --- | --- | --- |
| Boxing | 49 / 230 | 3.04  (2.17, 4.27) | 2.62  (1.68, 4.11) |
| Wrestling | 29 / 247 | 1.12  (0.74, 1.70) | 1.22  (0.73, 2.03) |
| Soccer | 34 / 248 | 1.54  (1.04, 2.26) | 1.23  (0.74, 2.06) |
| Other contact sports | 15 / 236 | 0.70  (0.41, 1.21) | 0.86  (0.47, 1.58) |
| Non-contact sports | 100 / 1,044 | 0.80  (0.61, 1.06) | 0.77  (0.54, 1.11) |
| General population (controls) | 106 / 1,386 | 1.0 (ref) | 1.0 (ref) |

SES, socioeconomic status. Co-morbidities are diabetes, hypertension, and

coronary heart disease. Lifestyle factors are alcohol intake, passing out due to

alcohol, body mass index, and smoking status.

**Supplemental Table 4. Hazard ratios (95% confidence intervals) for the association of former participation in contact sports with Alzheimer’s disease:**

**Correction for competing risk for cardiovascular disease in the Finnish cohort study**

|  | **Number of events / Number at risk** | **Age-adjustment**  (265 / 3391) | **Age-adjustment – with competing risk correction**  (265 / 3391) |
| --- | --- | --- | --- |
| Boxing | 46 / 230 | 4.45  (3.07, 6.45) | 4.15  (2.85, 6.04) |
| Wrestling | 31 / 247 | 1.90  (1.24, 2.90) | 2.12  (1.40, 3.21) |
| Soccer | 35 / 248 | 2.43  (1.62, 3.64) | 2.65  (1.77, 3.97) |
| Other contact sports | 12 / 236 | 0.85  (0.46, 1.56) | 1.01  (0.55, 1.85) |
| Non-contact sports | 70 / 1,044 | 0.89  (0.64, 1.23) | 1.07  (0.78, 1.49) |
| General population (controls) | 71 / 1,386 | 1.0 (ref) | 1.0 (ref) |

**Supplemental Table 5. Hazard ratios (95% confidence intervals) for the association of former participation in contact sports with Alzheimer’s disease:**

**Mutual adjustment for participation in other sports linked to Alzheimer’s disease in the Finnish cohort study**

|  | **Hazard ratios (95% confidence interval)** | | | |
| --- | --- | --- | --- | --- |
|  | **Age-adjustment – full cohort**  (188/1917 | **Age-, wrestling-, and soccer-adjustment**  (188 / 1917) | **Age-, boxing-, and soccer- adjustment**  (188 / 1917) | **Age-, boxing-, and wrestling-adjustment**  (188 / 1917) |
| Boxing | 4.00  (2.53,6.31) | 3.84  (2.41, 6.13) |  | - |
| Wrestling | 2.14  (1.32, 3.48) | - | 2.04  (1.24, 3.35) | - |
| Soccer | 1.98  (1.21, 3.24) | - | - | 1.97  (1.20, 3.22) |
| Other contact sports | 0.79  (0.40, 1.56) | - | - | - |
| Non-contact sports | 0.82  (0.56, 1.21) | - | - | - |
| General population (controls) | 1.0 (ref) | 1.0 (ref) | 1.0 (ref) | 1.0 (ref) |

In mutual-adjustment for sport participation, we covaried on those three sports shown to be related to

NDD outcomes in the present analyses. For instance, in analyses featuring boxing as the exposure of

interest, we controlled for wrestling and soccer.

**Supplemental table 6. Hazard ratios (95% confidence intervals) for the association of former participation in collision sports with Alzheimer’s disease:**

**Analyses of a non-missing dataset in the Finnish cohort study**

|  | **Number of events / Number at risk** | **Hazard ratios (95% confidence interval)** | |
| --- | --- | --- | --- |
|  |  | **Age-adjustment** | **Age-, SES-, co-morbidity-, and lifestyle factors-adjusted** |
| Boxing | 29/131 | 4.00  (2.54, 6.32) | 4.10  (2.55,6.61) |
| Wrestling | 24/132 | 2.14  (1.32, 3.48) | 2.11  (1.28, 3,48) |
| Soccer | 23/148 | 1.98  (1.21, 3.24) | 2.07  (1.23, 3.46) |
| Other collision sports | 10/170 | 0.79  (0.40, 1.56) | 0.80  (0.39,1.62) |
| Non-collision sports | 51/612 | 0.82  (0.56, 1.21) | 0.84  (0.55, 1.28) |
| General population (controls) | 51/724 | 1.0 (ref) | 1.0 (ref) |

**Supplemental box 1. Medline search terms**

"boxing"[MeSH Terms] OR "boxing"[All Fields] OR ("boxer"[All Fields] OR "boxer s"[All Fields] OR "boxers"[All Fields]) OR "martial"[All Fields] OR "wrestl*"[All Fields] OR ("football"[MeSH Terms] OR "football"[All Fields] OR "football s"[All Fields] OR "footballer"[All Fields] OR "footballer s"[All Fields] OR "footballers"[All Fields] OR "footballs"[All Fields]) OR ("soccer"[MeSH Terms] OR "soccer"[All Fields] OR "soccers"[All Fields]) OR ("hockey"[MeSH Terms] OR "hockey"[All Fields]) OR ("rugby"[MeSH Terms] OR "rugby"[All Fields]) OR ("racquet sports"[MeSH Terms] OR ("racquet"[All Fields] AND "sports"[All Fields]) OR "racquet sports"[All Fields] OR "lacrosse"[All Fields]) AND ("neurodegenerat*"[All Fields] OR "dement*"[All Fields] OR "alzheimer*"[All Fields]
